# A machine learning-based SNP-set analysis approach for identifying disease-associated susceptibility loci

**DOI:** 10.1101/2022.04.25.22274157

**Authors:** Princess P. Silva, Joverlyn D. Gaudillo, Julianne A. Vilela, Ranzivelle Marianne L. Roxas-Villanueva, Beatrice J. Tiangco, Mario R. Domingo, Jason R. Albia

## Abstract

**Introduction:** Identifying disease-associated susceptibility loci is one of the most pressing and crucial challenges in modeling complex diseases. Existing approaches to biomarker discovery are subject to several limitations including underpowered detection, neglect for variant interactions, and restrictive dependence on prior biological knowledge. Addressing these challenges necessitates more ingenious ways of approaching the “missing heritability” problem.

**Objectives:** This study aims to discover disease-associated susceptibility loci by augmenting previous genome-wide association study (GWAS) using the integration of random forest and cluster analysis.

**Methods:** The proposed integrated framework is applied to a hepatitis B virus surface antigen (HBsAg) seroclearance GWAS data. Multiple cluster analyses were performed on (1) single nucleotide polymorphisms (SNPs) considered significant by GWAS and (2) SNPs with the highest feature importance scores obtained using random forest. The resulting SNP-sets from the cluster analyses were subsequently tested for trait-association.

**Results:** Three susceptibility loci possibly associated with HBsAg seroclearance were identified: (1) SNP rs2399971, (2) gene LINC00578, and (3) locus 11p15. SNP rs2399971 is a biomarker reported in the literature to be significantly associated with HBsAg seroclearance in patients who had received antiviral treatment. The latter two loci are linked with diseases influenced by the presence of hepatitis B virus infection.

**Conclusion:** These findings demonstrate the potential of the proposed integrated framework in identifying disease-associated susceptibility loci. With further validation, results herein could aid in better understanding complex disease etiologies and provide inputs for a more advanced disease risk assessment for patients.

## Introduction

Understanding the emergence and progression of complex diseases incessantly pose challenges to researchers due to its intricate and multifactorial nature. These diseases are caused by interplays between genetics and environmental factors leading to a plethora of combinations that need to be considered in modeling. From the genetics’ aspect, understanding the etiology of complex diseases necessitates an extensive localization of significant genomic variations due to its polygenic nature [1, 2, 3]. Identifying these biomarkers, albeit elucidating only a portion of the entire underpinnings of complex diseases, could nevertheless aid in increasing patients’ chances of survival by allowing a more personalized and advanced disease risk assessment [4].

A genome-wide association study (GWAS) is the traditional approach employed to discover genetic biomarkers, i.e. single nucleotide polymorphisms (SNPs), associated with various traits and diseases [5]. GWAS has been successful in identifying several risk loci for a wide array of illnesses including cancer [6], Type 2 diabetes mellitus [7], Crohn’s disease [8], and coronary artery disease [9], among others. However, despite these achievements, GWAS faces limitations due to its individual-SNP analysis approach exacerbated by the high dimensionality of genomic datasets. As multitudinous individual association tests are performed, stringent thresholds must be adopted to account for error rates leading to underpowered detection [10]. This increases the probability of not detecting SNPs with small effects that are truly associated with a trait and could significantly contribute to phenotypic variability [11]. The traditional GWAS approach also fails to capture SNP-SNP interactions as it only tests for the marginal effects of SNPs and disregards the variants’ joint contributions to phenotypic expression. These interactions require explicit analysis since they are vital in addressing the “missing heritability” problem [12] which states that single genetic variations are insufficient in explaining the entire heritability of a trait.

Under the “polygenic paradigm”, refining statistical models, such as increasing sample sizes [13] and reducing the number of tests employed [14], is crucial in increasing the chances of discovering true associations. Empirical evidence [15, 16] has shown that as sample size increases, GWAS continues to yield more novel trait-associated loci. However, this approach is not always feasible [14] especially for studies involving small populations and diseases with low prevalence. For this reason, it is more viable to reduce the number of tests employed to relax the stringent conditions used to consider genomic variants as significant. Existing approaches to this latter strategy include haplotype-based association analysis and SNP-set analysis, both of which also address the inability of GWAS to capture SNP-SNP interactions [17, 18]. Haplotype-based analysis [19] accounts for linkage disequilibrium between SNPs; while SNP-set analysis, e.g. gene-based [20] and pathway-based analyses [21], considers the joint effects of variants on phenotypic expression. Aside from addressing the aforementioned GWAS’ limitations, SNP-set analysis further permits hypothesis testing on associations possibly existing between wider loci and traits [18]. However, when this type of analysis groups SNPs based on prior biological knowledge, a study’s success may be hampered when information on genetic variations and competitive pathways related to the trait are insufficient. To allow a less restricted analysis, it is necessary to explore other methods of forming SNP-sets using information independent of a priori biological knowledge.

Machine learning (ML) is an innovative and powerful approach used in solving complex problems in various fields and disciplines due to its capability to handle and analyze high-dimensional datasets [22, 23, 24]. Several studies have already demonstrated the usability of ML in genomic datasets [25, 26, 27]; however, to our knowledge, there is only a handful of existing literature discussing its application to SNP-set formation [28, 29, 30, 31]. These studies employed cluster analysis to form SNP-sets in a data-driven manner. This approach could subsequently lead to the identification of novel risk loci associated with a trait [31], albeit there may be problems related to computational complexity and cost. As genomic datasets are usually of high dimension, it is susceptible to the “curse of dimensionality” [32, 33], a problem that could be addressed by solely clustering the SNPs found in certain genomic regions that are known to play a role in trait development [29, 30]. However, this approach defeats the purpose of performing an inclusive analysis as the search for significant biomarkers is restricted by relatively narrow regions. For a more varied selection of SNPs to analyze, dimensionality reduction techniques based on random forest (RF) could be used to reduce dataset dimensions before conducting cluster analysis. RF has been widely incorporated in SNP research [25, 34, 35, 36] due to its significant properties: (1) a nonparametric nature that allows the establishment of predictive models without the need for preliminary statistical assumptions, and (2) the capability to provide an importance score, i.e. variable importance measure (VIM) for each SNP, which increases the probability of detecting highly relevant biomarkers.

Cluster analysis and random forest have already been proven applicable and effective in genomic data analysis, specifically in identifying predictive and presumably disease-associated SNPs [31, 37]. However, based on the literature review, the integration of these approaches has not been explored on SNP data. This study aims to incorporate these two techniques to augment previous GWAS findings and allow the discovery of novel trait-associated susceptibility loci. The study implements the proposed integrated framework using the following three-step algorithm:

1. Dimensionality reduction through RF;
2. SNP-set formation through cluster analysis involving top-ranking SNPs from Step 1 and SNPs considered by GWAS to be significantly associated with the trait of interest (termed in this study as ‘GWAS-identified SNPs’); and
3. Association testing on the resulting SNP-sets from Step 2.

In Step 1, dimension reduction is implemented using random forest feature selection to circumvent the “curse of dimensionality” problem associated with analyzing high-dimensional SNP datasets [35]. In Step 2, top-ranking SNPs determined from the results of Step 1 and GWAS-identified SNPs are subjected to cluster analysis to evaluate shared similarities among the variants and form SNP-sets. Finally, Step 3 involves testing the SNP-sets derived from Step 2 for trait-association. The proposed methodology was applied to the GWAS data by [39] wherein the phenotype of interest is hepatitis B virus surface antigen (HBsAg) seroclearance, a marker for clearance of chronic hepatitis B virus (HBV) infection.

## Methodology

This study proposes a novel machine learning-based SNP-set analysis approach for identifying disease-associated susceptibility loci. RF, cluster analysis, and previous GWAS findings were integrated into a single framework to increase detection power and account for SNP-SNP interactions—factors that are vital in addressing the “missing heritability” problem. The entire analysis is divided into three main parts: dimension reduction, SNP-set formation, and association testing. Fig. 1 shows the architecture of the proposed integrated framework.

**Fig. 1:**
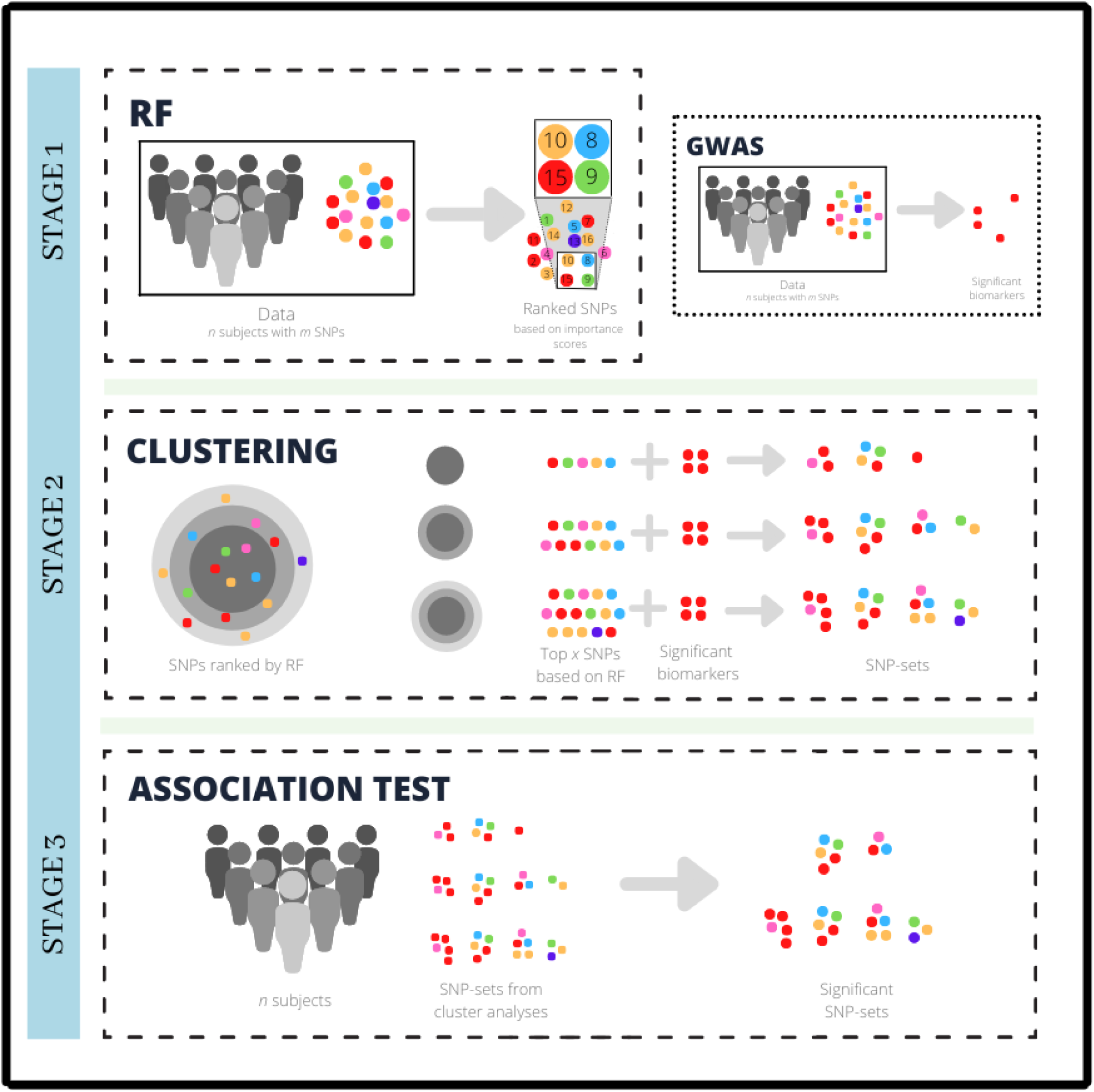
The architecture of the proposed integrated framework. In Stage 2, SNPs in concentric circles in darker shades of gray represent higher-ranking SNPs based on RF.

### Data Description and Preprocessing

The data used in this study was adopted from the GWAS conducted by [39] which aimed to identify susceptibility loci associated with HBsAg seroclearance among patients with chronic hepatitis B. The dataset is composed of 1,365,088 SNPs collected from 200 subjects of Korean ethnicity. The subjects were further divided into two groups: the cases (n = 100), which consist of patients who had experienced HBsAg seroclearance before the age of 60, and the controls (n = 100) comprising of patients who exhibited high levels (> 1000 IU/mL) of HBsAg at ≥ 60 years of age. An additive genetic model was utilized to transform the SNP dataset wherein 0, 1, and 2 were used to represent homozygous dominant, heterozygous, and homozygous recessive, respectively.

### Dimension Reduction

Dimension reduction is commonly a prerequisite in analyzing SNP datasets as large amounts of features exceed the capability of analytical approaches in performing fast and effective analyses. In this study, VIM of RF was used to reduce dataset dimension by identifying highly predictive and informative SNPs prior to conducting cluster analysis. RF has been widely utilized in analyzing SNP data primarily due to its capacity to build a predictive model without making any assumptions about the underlying relationship between genotype and phenotype [40]. In RF, the predictive abilities of multiple decision trees, which are trained on bootstrap samples of the data, are consolidated to generate the final output prediction. In addition, randomization is not only induced by bootstrapping but also introduced at the node level when growing a tree. It selects a random subset of SNPs at each node of the tree as candidates to find the best split for the node. In estimating the importance of SNPs, RF calculates the Gini importance which quantifies the difference between a node’s impurity and the weighted sum of the impurities of the two descendent nodes.

Mathematically, the importance of *SNP_j_* is determined by summing the decrease in impurity (*ΔI*) for all the nodes *t*, where *SNP_j_* is split. The decreases in impurity are weighted by fractions of samples in the nodes *p*(*t*) and averaged over all trees in the forest. The Gini variable importance is then given by,

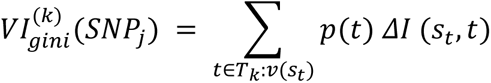

where *T_k_* is the number of nodes in the *k^th^* tree, 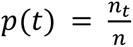 is the fraction of the samples reaching node *t*; and *v*(*s_t_*) is the variable used in the split *s_t_*.

Step 1 of the proposed integrated framework uses a random forest classifier that is initially trained on the dataset and evaluated using leave-one-out cross-validation (LOOCV). LOOCV uses *N* – *1* observations as the training set and the excluded observation as the testing set, where *N* is the number of samples. This ensures reliability and unbiasedness in the estimation of model performance. The final feature importance score of a SNP is then calculated by averaging the scores of the said SNP obtained by RF for every fold in LOOCV.

### SNP-set formation

This study exploited the similarities shared among SNPs to identify novel susceptibility loci associated with HBsAg seroclearance. The analysis utilized the unsupervised machine learning method known as cluster analysis which aims to separate data points into distinct groups such that more similarities are shared among objects within the same group than objects belonging to different groups. Similarities between SNPs can be quantified in terms of *agreement,* i.e. based on the occurrence of sequence alterations computed via matching coefficients and measures of correlation, or *dependence*, i.e. based on the presence or absence of dependence quantified via measures based on the χ2-statistic [41]. This study adopts an *agreement*-based similarity measure by employing the method proposed in [30]. This method modified an agglomerative hierarchical clustering algorithm with average linkage for continuous data to develop a Hamming distance-based algorithm for determining SNP-sets. Hamming distance is a similarity measure used to calculate the number of dissimilar components between two categorical data points of the same size [42]. Applied to SNP data, the Hamming Distance *d^HAD^* between SNPs *i* and *j* would be,

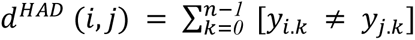

where *n* is the total number of subjects and *y_k_* is the genotype of the *k*th subject. The similarity measure was adapted on SNP datasets based on the premise that the more individuals carrying the same genotype concerning two given SNPs or two SNP-sets (signified by a relatively small Hamming distance), the more similar the variants are and more likely to cluster [30].

Multiple cluster analyses were performed exclusively on GWAS-identified and top-ranking SNPs obtained by random forest. As shown in Table 1, the number of SNPs analyzed was gradually increased to achieve a higher likelihood of discovering novel susceptibility loci. Each implementation resulted in candidate SNP-sets identified using the following parameters: *percentile cut* which specifies the height wherein a dendrogram will be cut and *minimum cluster size* which dictates the minimum number of SNPs for all clusters.

**Table 1.** Number of SNPs subjected to cluster analysis

| Cluster analysis | Number of SNPs included* |
| --- | --- |
| 1 | 1047 |
| 2 | 2044 |
| 3 | 3041 |
| 4 | 4038 |
| 5 | 5036 |

\*The set of SNPs included in the cluster analysis is the union of the 52 significant SNPs from Kim et. al.'s GWAS [39] and the top biomarkers identified by random forest (starting from top 1000 to top 5000 SNPs in increments of 1000).

### Association test

Hamming distance-based association tests (HDAT) [30] were employed to identify the candidate SNP-sets significantly associated with HBsAg seroclearance. The presence of association depends on the amount of difference in the biomarkers found in cases and controls. Minor alleles were incorporated in the equations as it reveals more similarities in the genomes of two individuals than common alleles [43]. A comprehensive discussion of the equations used in HDAT can be found in [30]. Permutation test, a non-parametric test used to evaluate the statistical significance of a model through randomization, is used to compute the p-value of each SNP-set. The test calculatesthe *p-value* by permuting the dataset and constructing a test-statistic distribution and evaluating the probability that a test-statistic would be equal to or more extreme than the initial computed value.

## Results

### Top-ranking SNPs from dimension reduction

This study used random forest feature selection to reduce dataset dimensions prior to conducting cluster analysis. Specifically, random forest was employed to rank SNPs based on their feature importance score, a measure which determines a variant’s relevance in making accurate phenotype predictions. SNPs are assigned a feature importance score based on the average scores for every fold in LOOCV to eliminate bias and ensure robustness. Investigation into the functional significance of three of the top five biomarkers ranked by RF led to possible connections between the variants and HBsAg seroclearance. SNPs rs28588178 (top-ranking SNP), rs1994209 (3rd-ranking SNP), and rs7958186 (5th-ranking SNP) are linked with Cadherin 4 (CDH4), PIG11, and PCED1B, respectively—genes reported to be associated with hepatocellular carcinoma (HCC) [44, 45, 46], a disease that can develop due to the presence of the hepatitis B virus.

### Generated SNP-sets

Upon performing multiple cluster analyses, a total of 108 candidate SNP-sets were identified at a percentile cut of 0.9 and a minimum cluster size of 3. SNP-sets with the maximum number of SNPs were chosen in cases where there were overlaps to maximize the information obtained from the analyses.

SNP-sets containing SNPs which were considered significant in a previous GWAS were investigated as the variants sharing high degrees of similarity with GWAS-identified SNPs may also provide insights into trait etiology. As shown in Table 2, SNPs rs2399971, rs2119977, rs6826277, rs35689347, rs1505687, and rs741229 were grouped with at least one of the variants reported to be significantly associated with HBsAg seroclearance. No information regarding possible association existing between the latter five SNPs and the phenotype of interest was found; meanwhile, the opposite was true for rs2399971. Notably, albeit rs2399971 had not reached the cut-off value used in the GWAS performed by Kim et al. [39] on the whole study population, it was nevertheless found to be significantly associated with HBsAg seroclearance in the subjects who had received antiviral treatment [39]. Fig. 2 shows the dendrogram of the GWAS-identified SNPs together with the aforementioned six variants and as presented, the SNPs belonging to the SNP-set which contains rs2399971 shows the least height differences, indicating that the SNPs in the set are more similar to each other than the variants found in other clusters.

**Fig. 2:**
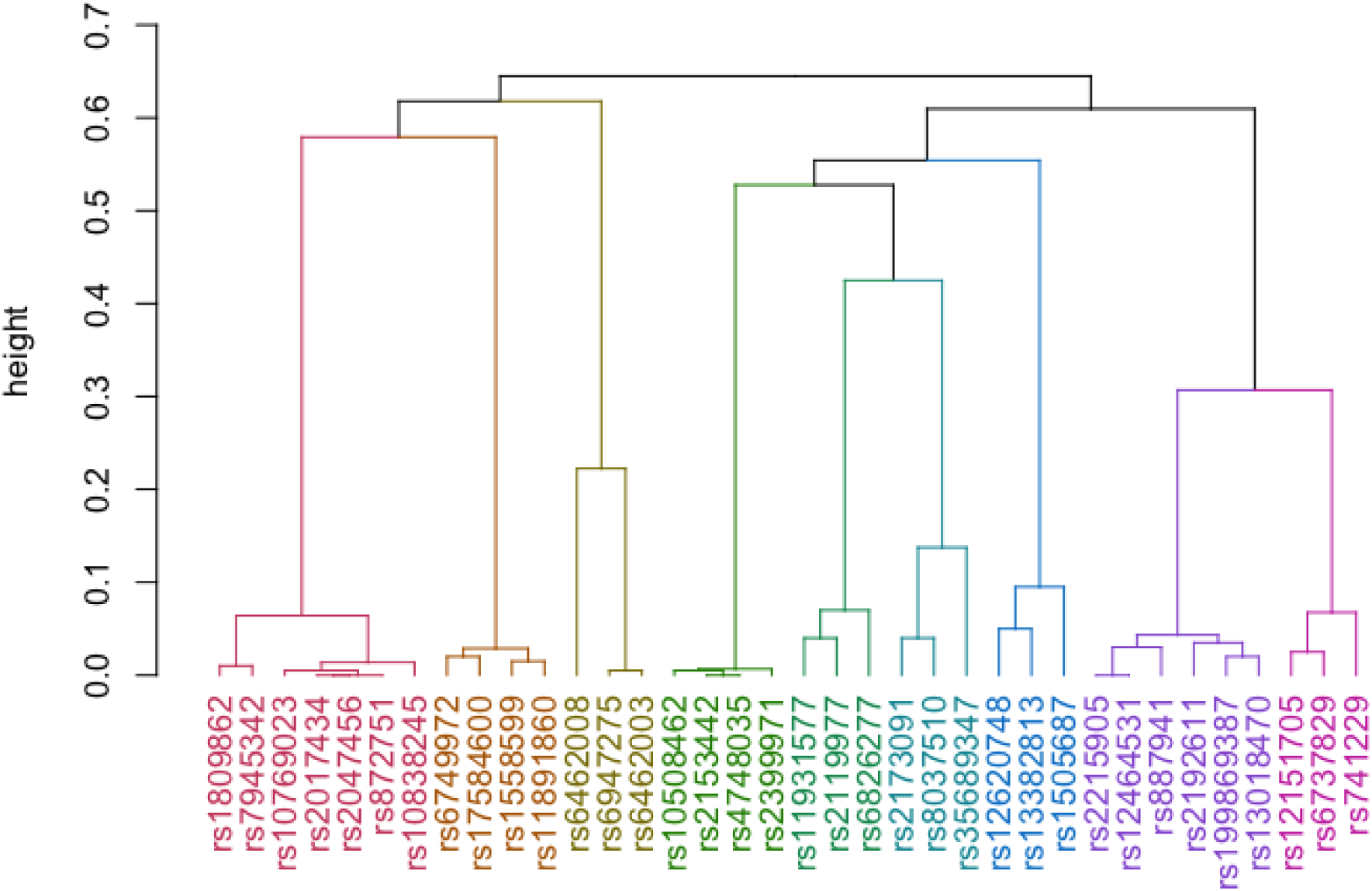
Dendrogram of the SNPs listed in Table 2.

**Table 2.**
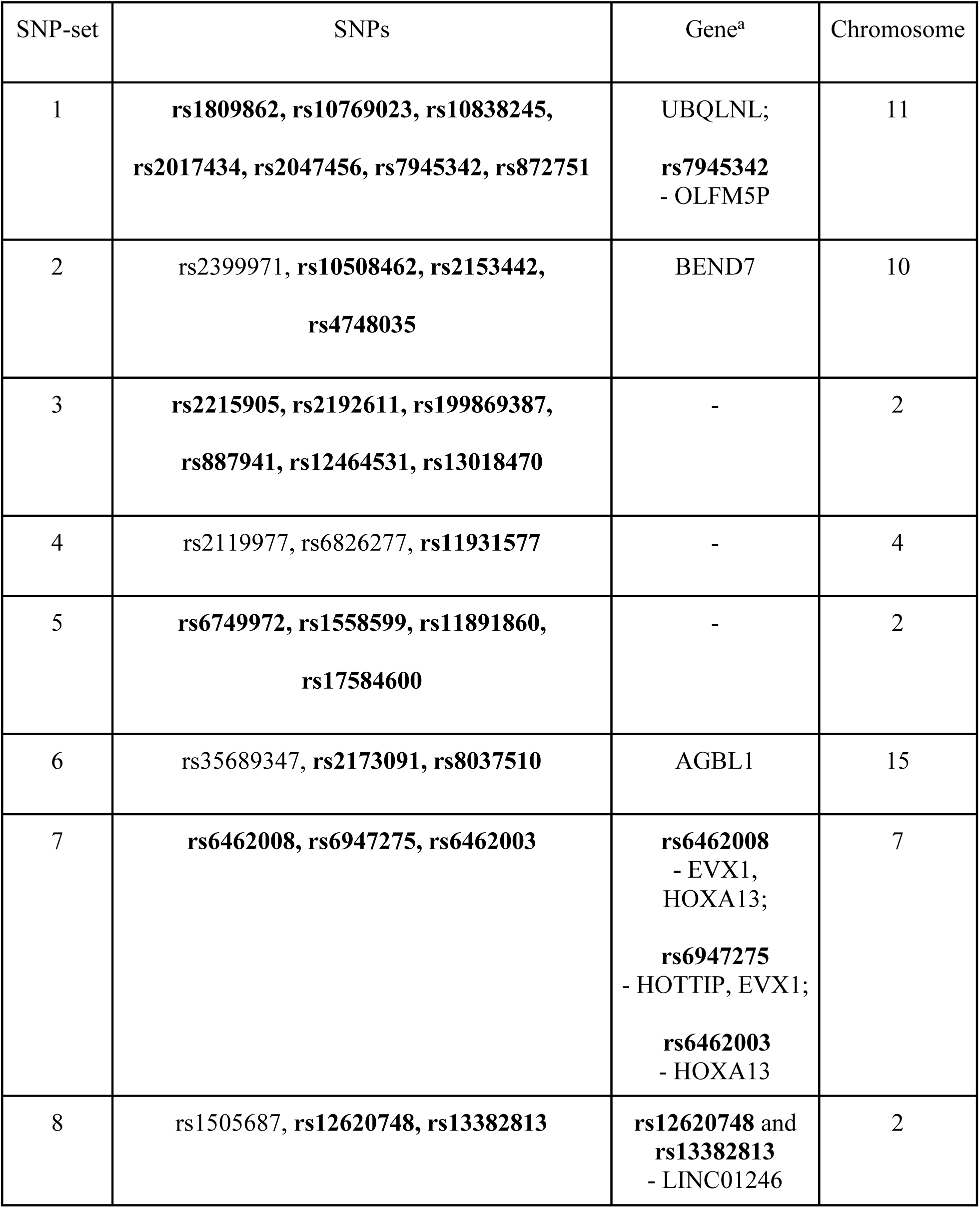
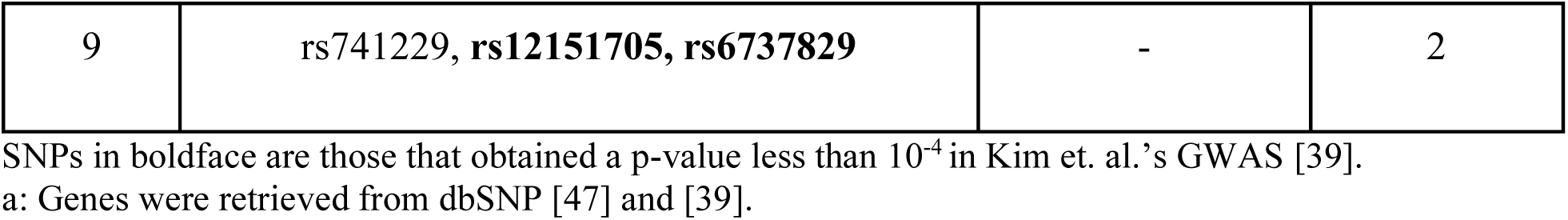
Cluster memberships of the SNPs that obtained a p-value less than 10^-4^ in Kim et. al.’s GWAS [39]

### Significant SNP-sets

Hamming distance-based association test (HDAT) was performed on the candidate SNP-sets to further identify SNPs possibly associated with HBsAg seroclearance. After performing a Bonferroni correction for multiple tests, 11 SNP-sets significantly associated with HBsAg seroclearance (*p-value* < 0.0005) were identified, the majority of which (7 out of 11) were found to harbor at least one of the GWAS-identified SNPs. Among the SNP-sets obtaining the lowest p-values, the set which obtained the highest test statistic is the one composed of rs1809862, rs10769023, rs10838245, rs2017434, rs2047456, rs7945342, and rs872751—all GWAS-identified SNPs [39]. All these variants reside in 11p15.4, a region that shows a possible correlation with HBsAg seroclearance. In a study by [48], it was observed that among hepatocellular carcinoma cases, more than 20 percent loss of heterozygosity (LOH) was shown for locus 11p, wherein region 11p15 was commonly affected. Moreover, a significant correlation was found to exist between LOH on 11p and HBsAg positivity. Specifically, results showed that there is a significantly higher frequency of LOH on 11p among hepatitis B virus carriers [48].

Table 3 shows the five significant SNP-sets which do not hold any of the GWAS-identified SNPs. No supporting evidence was found regarding possible associations between the individual variants belonging to the five SNP-sets and HBsAg seroclearance. Nonetheless, interesting findings were discovered when SNPs were analyzed collectively. Results showed that three out of the five SNP-sets in Table 3 harbor SNPs residing in similar genes, i.e. there is a corresponding gene for each distinct set. These are the following: (1) LOC105373438 for SNP-set 3, (2) LINC00578 for SNP-set 4, and (3) STOX2 for SNP-set 5. In [49], LINC00578 was reported to be a prognostic marker for pancreatic cancer (PC), a disease for which hepatitis B has been suggested to be a risk factor [50, 51, 52], increasing the likelihood of PC by 24% [53].

**Table 3.** SNP-sets obtaining the lowest p-values (excluding those that harbor variants reported by Kim et al. [39] to be significantly associated with HBsAg seroclearance)

| SNP-set | List of SNPs | p-value* |
| --- | --- | --- |
| 1 | rs6731235, rs199703414, rs16829541, rs1485096,<br>rs2341849 | 0.0002 |
| 2 | rs28365850, rs62625038, rs17102970 | 0.0004 |
| 3 | rs59659073, rs10754962, rs2380525 | 0.0004 |
| 4 | rs200957040, rs1499880, rs4857702 | 0.0004 |
| 5 | rs12644266, rs13130260, rs6815422 | 0.0001 |

\*p-values were obtained from 10000 permutations

## Discussion

This study aims to discover novel trait-associated susceptibility loci by augmenting previous GWAS findings using a machine learning-based SNP-set analysis approach built on the integration of RF and cluster analysis. Investigation into the functional relevance of variants found in the same SNP-set containing GWAS-identified SNPs and SNP-sets obtaining significant p-values led to the discovery of loci that may also contribute to phenotypic expression yet overlooked by GWAS as a consequence of stringent detection conditions and neglect for SNP-SNP interactions in the experimental design. The novelty in our proposed method lies in the GWAS-based and data-driven approach in feature selection prior to cluster analyses. This study did not restrict the discovery of susceptibility loci to a certain genomic region alone as the criteria for selecting SNPs depend on statistical significance and predictive powers. As a result, the resulting SNP-sets implicated a varied selection of genes and cytobands.

The proposed method was applied on an HBsAg seroclearance GWAS data [39] and was able to detect SNP rs2399971 as it showed a high degree of similarity with GWAS-identified SNPs. Note that variant rs2399971, albeit not considered significant in the GWAS conducted on the whole study population (obtaining a p-value of 1.05×10^-4^ wherein the cut-off p-value used was 1.00×10^-4^), nevertheless exhibited significance in the subgroup analysis performed (p-value of 4.60×10^-5^). This result demonstrates that by reducing the unit of analysis into groups and exploiting previous GWAS findings, an increase in detection power could be achieved as a result of pooled strengths of signal. Through SNP-set analysis, it also becomes possible to generate hypotheses not only on SNPs but also on other larger biological units such as genes or cytobands [29, 18]. For instance, gene LINC00578 and locus 11p15, regions implicated by two of the SNP-sets with the lowest p-values, have shown potential in understanding HBsAg seroclearance as both are linked with diseases associated with the presence of hepatitis B virus infection. By mapping out these implicated regions and identifying shared susceptibility loci with a well-researched phenotype, a better understanding of the intricate underpinnings of the trait of interest could be achieved. For instance, some of the SNPs associated with height may be considered in understanding the etiology of HBsAg seroclearance as 11p15 has been reported to harbor genes responsible for growth and development [54]. Furthermore, elevations in alanine transaminase (ALT) level, a consideration in declaring HBsAg seroclearance, was found to be an important factor for growth impairment in children [55].

Despite the advantages, the proposed method is subject to several limitations such as time and computational constraints affecting the total number of SNPs for inclusion in the cluster analyses; therefore, variants possibly associated with the trait but obtaining low feature importance scores might not be accounted for. Secondly, parameter values would still have to be tuned by utilizing specific measures such as gap statistics [56, 57] to ensure an optimal number and a more cohesive composition of SNP-sets. Lastly, there is a lack of previous research on the integration of unsupervised and supervised machine learning techniques in analyzing SNP data as well as a scarcity of studies on SNP-set formation and trait-association. Considering these limitations, results obtained from the analysis necessitate further biological investigation.

## Conclusion

This study aims to identify disease-associated susceptibility loci by augmenting previous GWAS findings using the integration of RF and cluster analysis. The proposed approach was applied to a hepatitis B virus surface antigen (HBsAg) seroclearance GWAS data [39]. Thereafter, the researchers were able to detect rs2399971, a variant that was not considered to be significantly associated with the phenotype in the main GWAS, but which obtained a significantly low p-value in a subgroup analysis [39]. Results of the association tests conducted on the generated SNP-sets led to the implication of gene LINC00578 and locus 11p15. The former was linked with pancreatic cancer [49] and the latter with hepatocellular carcinoma [48], diseases associated with hepatitis B virus infection. Researchers who aim to extend this study could experiment on different supervised learning techniques for feature selection and utilize other similarity measures for clustering SNPs. With further investigation and validation, insights gleaned using the proposed framework could also be integrated into prediction models to aid in quantifying patients’ risks for trait or disease development.

## Data Availability

All data produced in the present study are available upon reasonable request to the authors

## Compliance with Ethics Requirement

This study used the data provided in [39] which was a project approved by the ethics committees at Korea University Anam Hospital (ED13220) and conducted in agreement with the ethical principles of the Declaration of Helsinki. According to the project’s ethical declaration, all patients provided written informed consent for participation and use of their data for research purposes.

## CRediT authorship contribution statement

**Princess P. Silva:** Conceptualization, Methodology, Software, Validation, Formal Analysis, Investigation, Writing – Original Draft, Writing – Review and Editing, Visualization. **Joverlyn D. Gaudillo:** Conceptualization, Methodology, Validation, Investigation, Writing Original Draft, Writing - Review and Editing, Supervision, Data Curation. **Julianne A. Vilela:** Conceptualization, Writing – Review and Editing. **Ranzivelle Marianne L. Roxas-Villanueva:** Resources, Writing - Review and Editing, Project Administration, Funding Acquisition. **Beatrice J. Tiangco**: Resources, Project Administration, Funding Acquisition. **Mario R. Domingo:** Resources, Project Administration, Funding Acquisition. **Jason R. Albia:** Resources, Writing – Review and Editing, Project Administration, Funding Acquisition.

## Acknowledgment

The authors would like to thank the Department of Science and Technology – Philippine Council for Health Research and Development (DOST-PCHRD) for providing the necessary funding and assistance that made this study possible. The analysis conducted herein acts as a preliminary study for the project sponsored by DOST-PCHRD entitled “AI-driven Integration of Genomic, Ultrasound, Serum Biomarkers, and Clinical data for Early diagnosis of Liver Cancer” under the program “Early CANcer Detection in the LivEr of Filipinos with Chronic Hepatitis B Using AI-Driven Integration of Clinical and Genomic Biomarkers (CANDLE Study)”.

## Appendix

**Table A.**
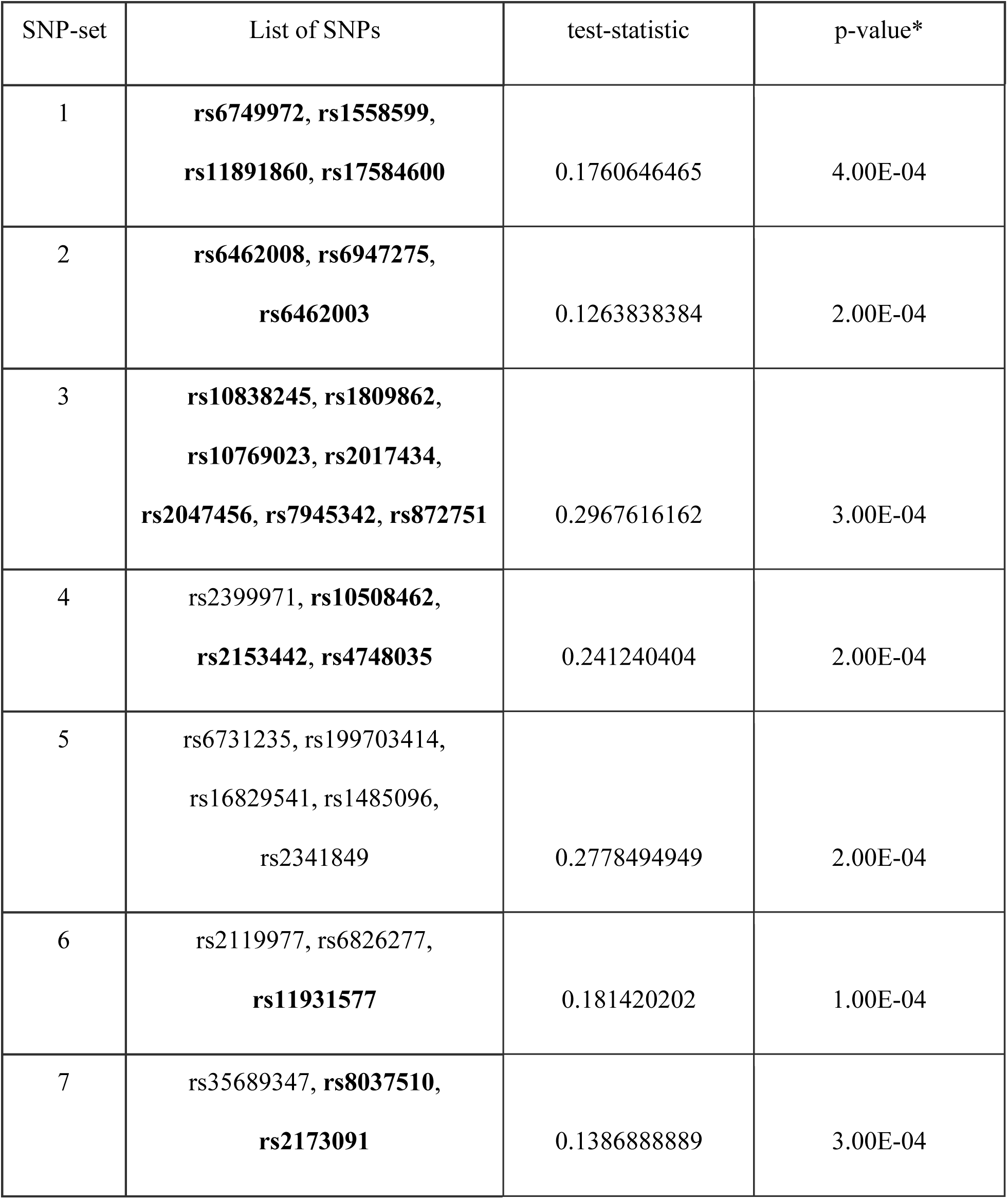
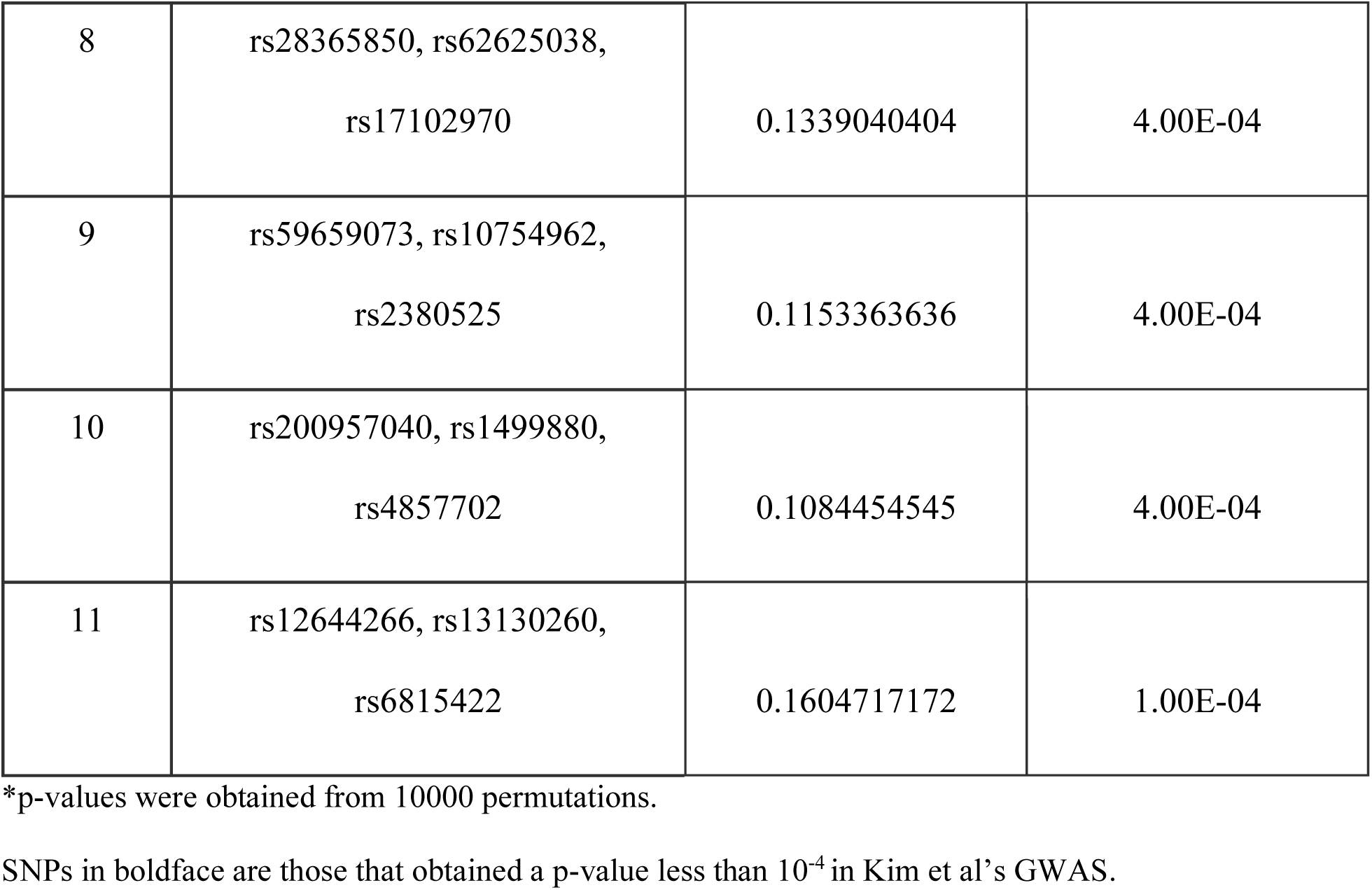
SNP-sets significantly associated with HBsAg seroclearance (*p-value* < 0.0005)

| SNP-set | List of SNPs | test-statistic | p-value* |
| --- | --- | --- | --- |
| 1 | **rs6749972, rs1558599,**<br>**rs11891860, rs17584600** | 0.1760646465 | 4.00E-04 |
| 2 | **rs6462008, rs6947275,**<br>**rs6462003** | 0.1263838384 | 2.00E-04 |
| 3 | **rs10838245, rs1809862,**<br>**rs10769023, rs2017434,**<br>**rs2047456, rs7945342, rs872751** | 0.2967616162 | 3.00E-04 |
| 4 | rs2399971, **rs10508462,**<br>**rs2153442, rs4748035** | 0.241240404 | 2.00E-04 |
| 5 | rs6731235, rs199703414,<br>rs16829541, rs1485096,<br>rs2341849 | 0.2778494949 | 2.00E-04 |
| 6 | rs2119977, rs6826277,<br>**rs11931577** | 0.181420202 | 1.00E-04 |
| 7 | rs35689347, **rs8037510,**<br>**rs2173091** | 0.1386888889 | 3.00E-04 |
| 8 | rs28365850, rs62625038,<br>rs17102970 | 0.1339040404 | 4.00E-04 |
| 9 | rs59659073, rs10754962,<br>rs2380525 | 0.1153363636 | 4.00E-04 |
| 10 | rs200957040, rs1499880,<br>rs4857702 | 0.1084454545 | 4.00E-04 |
| 11 | rs12644266, rs13130260,<br>rs6815422 | 0.1604717172 | 1.00E-04 |

\*p-values were obtained from 10000 permutations.
SNPs in boldface are those that obtained a p-value less than $10^{-4}$ in Kim et al's GWAS.

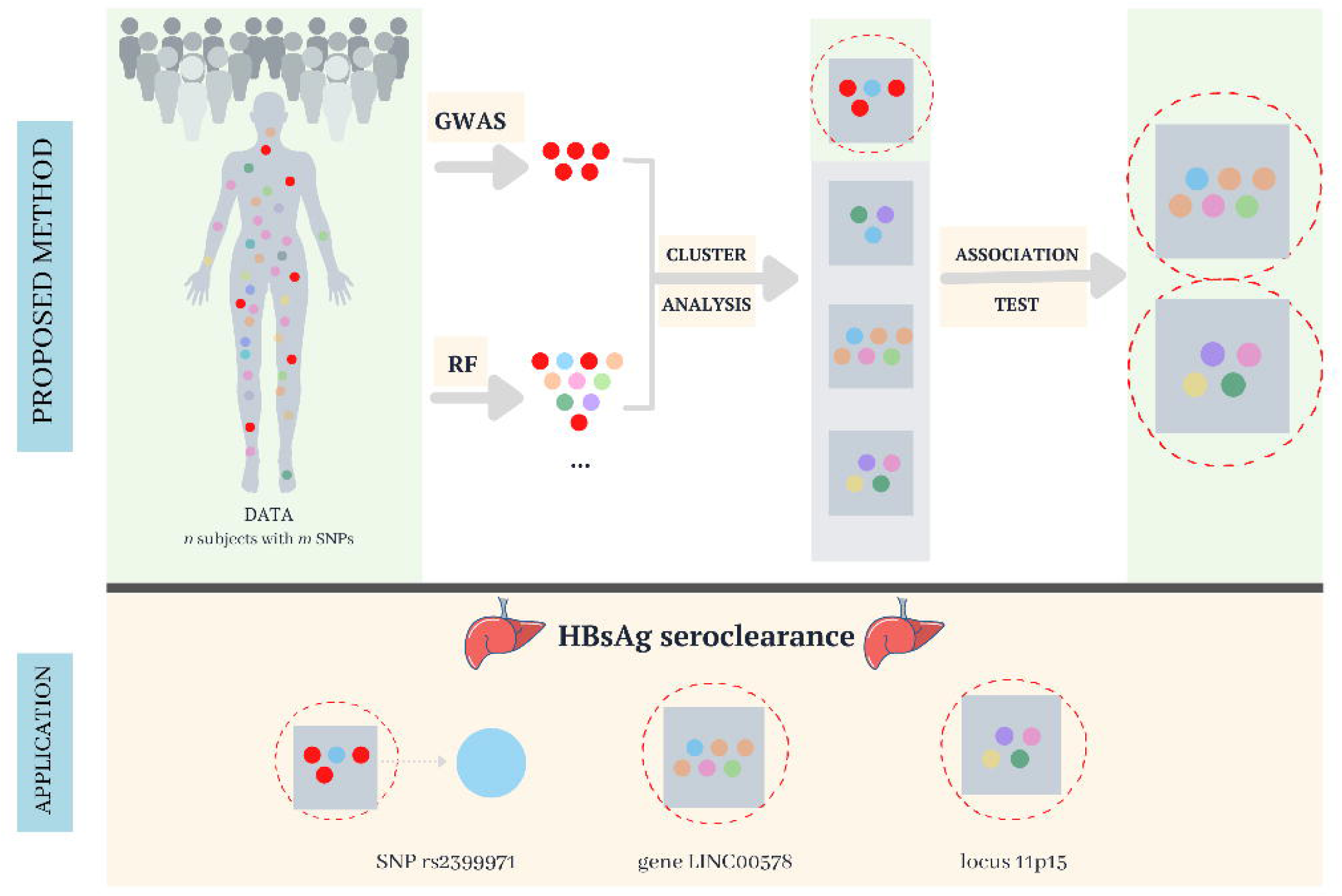

